# Capping Mobility to Control COVID-19: A Collision-based Infectious Disease Transmission Model

**DOI:** 10.1101/2020.07.25.20162016

**Authors:** Yunfeng Shi, Xuegang Ban

## Abstract

We developed a mobility-informed disease-transmission model for COVID-19, inspired by collision theory in gas-phase chemistry. This simple kinetic model leads to a closed-form infectious population as a function of time and cumulative mobility. This model uses fatality data from Johns Hopkins to infer the infectious population in the past, and mobility data from Google, without social-distancing policy, geological or demographic inputs. It was found that the model appears to be valid for twenty hardest hit counties in the United States. Based on this model, the number of infected people grows (shrinks) exponentially once the relative mobility exceeds (falls below) a critical value (∼30% for New York City and ∼60% for all other counties, relative to a median mobility from January 3 to February 6, 2020). A simple mobility cap can be used by government at different levels to control COVID-19 transmission in reopening or imposing another shutdown.

## Introduction

Novel Coronavirus Disease 2019 (COVID-19) pandemic is probably the most challenging issue facing the world today. The loss of lives, the emerging new social norms and the stagnant if not collapsing economy are hallmarks of an era that the world has not seen in recent memory. COVID-19 has already exacerbated many pressing issues such as food, poverty, social justice, health care, education, government accountability, with a possible singular exception of pollution. There are tremendous economic, social and political pressures to reopen society. Thus, it becomes critical to have quantitative forecast of COVID-19 infections and fatalities to guide federal, state and local governments in formulating reopening and resource allocation policy.

The classical model for infectious disease transmission is the SIR (susceptible, infectious, recovered) model by Kermack and Mckendrick,^1^ a kinematic compartment model considering the change of infectious population, susceptible population and recovered (immune) population. According to the SIR model, a disease outbreak will occur once the basic reproduction number *R*_*0*_ (the number of new infections in a completely susceptible population prior to removal) is higher than the reciprocal of the initial susceptibility of the population (fraction of people without immunity). The idea of “herd immunity” is essentially to control the disease outbreak by allowing infection to immunize the population thus reducing the susceptibility. The original SIR model can be expanded to include immunity with limited lifetime,^2,3^ vital dynamics with birth and death,^4^ maternally-derived immunity,^5^ or incorporate an extra population for exposed individuals not yet infectious.^6^ In addition, statistical tools have been frequently used to describe the probabilistic nature of disease transmission, uncertainty in the disease data and variations in pathogens to provide better prediction.^7–9^ Using such advanced compartment models, disease spread dynamics can be predicted by COVID-19 Simulator,^10^ Delphi,^11^ and LMIC.^12,13^

Stay-at-home order (or the more draconian city-wide lockdown) and other public health intervention measures (e.g., social distancing) have been proven the most effective ways to combat COVID-19 outbreaks. Reduced mobility leads to reduced contacts and disease transmission. It has been shown that there is clear correlation between mobility patterns and COVID-19 transmission.^14^ Basellini et al. have linked excess mortality to human mobility in England and Wales.^15^ Vollmer et al. estimated disease transmission using mobility in Italy.^16^ The effect of travel restriction policies on the global COVID-19 outbreak has also been studied by Chinazzi et al. using a metapopulation disease transmission model.^17^ The stay-at-home policies could lead to the so-called “suppressed equilibrium”, and possible eventual “heard-immunity” yet with less overall death. To this end, Tung and co-workers have built a statistical model to account for diminished infectivity due to social distancing.^18^ Numerical simulations of stochastic disease spreading also consider mobility directly by incorporating the sociodemographic and population mobility data (such as the GLEAM framework^19^). LMIC also incorporates mobility data by dynamically varying the reproductive number using Google mobility data.^13^

There are probably close to a hundred different COVID-19 models available with varying predictability. To our best knowledge, there is no mechanistic model that incorporates mobility and has a close-formed solution, without which it is difficult to evaluate the efficacy of the model and avoid ever-growing number of parameters causing overfitting. Here, we introduce a minimalist’ s mechanistic infectious disease model incorporating mobility information directly, motivated by molecular collision used in gas-phase chemical reaction. The key of this model is to acknowledge that the total contacts of a community are proportional to the cumulative mobility, which should play the role of time in epidemic outbreaks. This simple model has an analytical solution relative to a reference date, which can be directly tested against publicly available data. It was found that infections in twenty counties in the United States with the highest fatalities as of July 12^th^, 2020 agree well with the model. Importantly, we found there exists a critical value for the relative mobility that dictates whether outbreak will occur, which could be utilized in designing reopening policy or possibly imposing a second shutdown.

### Mobility-informed Disease-Transmission Model

The fundamental process in understanding the chemical reaction in gas-phase is the binary collisions between molecules. As shown in Figure 1(a), the green molecule travels along a path that could encounter other molecules resulting in collisions and possibly reactions. Analogously, a person travels along a path that could encounter other people or contaminated surfaces/air, and possibly gets infected. Thus, for a given duration, the shorter the path a molecule covers (equivalently, the fewer number of places a person visits), the lower the number of collisions (equivalently, the lower number of encounters leading to infections). It should be noted that the original SIR model is in fact motivated by McKendrick’ s “particle collisions” metaphor,^9^ in which the mobility information is not explicitly considered.

**Figure 1.**
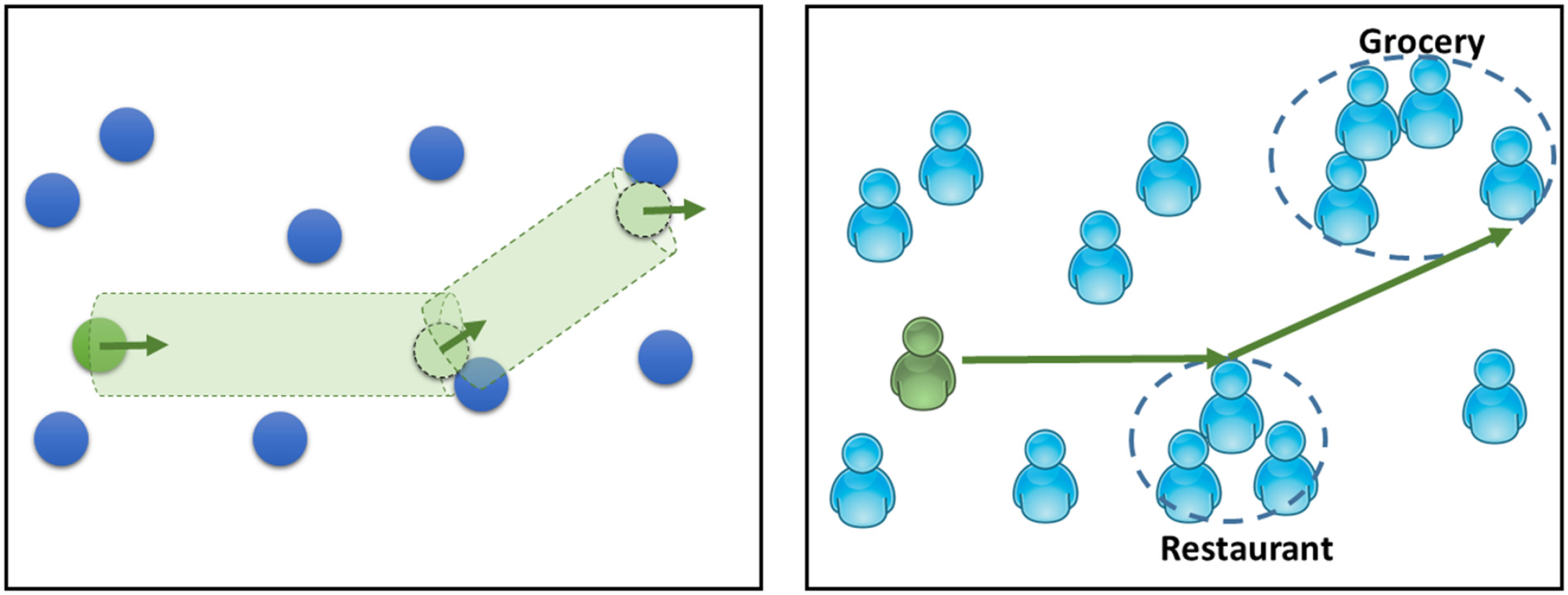
A comparison between binary collisions of gas-phase molecules (left) and the encounters between people (right). As the number of collisions per time is proportional to velocity of the molecule, the number of human encounters per time is proportional to the mobility of the person.

The overarching assumption of this mobility-informed disease-transmission model is that, the number of encounters of a person during a given time is proportional to his or her mobility *m*. Mobility here is defined as the number of visits to activity locations within a day, relative to that in the baseline day before the pandemic. Due to the convenient linear relation, this assumption can be easily extended to a community such that the total number of encounters is proportional to the sum of the community mobility (the number of people times the average mobility). In addition, the infection must be also proportional to the fraction of the population that is currently infectious, as in the SIR model. For simplicity, this model does not consider the immunity of recovered population, which is reasonable as the overall infected population is still low and the short duration of immunity after infection.^20^ The total population is assumed to be a constant without considering birth or death. Similar to the SIR model, recovery is proportional to the current infectious population, we have,

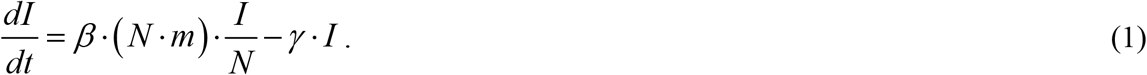

*I* is the current infectious population. *N* is the total population. *I/N* is the fraction of population that is currently infectious. *m* is the current mobility. *β* is the community-specific infection rate. *γ* is the recovery rate. Here we assume both *β* and *γ* are constant for simplicity. Integrate from a reference time *t*_*0*_, we have,

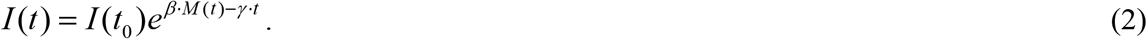

Here 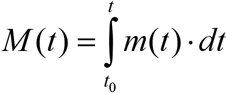 is the cumulative mobility from *t*_*0*_ to *t*. The rather simple two-parameter analytical solution to this mobility-informed disease transmission model permits critical examination. A direct test can be conducted by recasting the infectious population *I(t)* in Eq. (2),

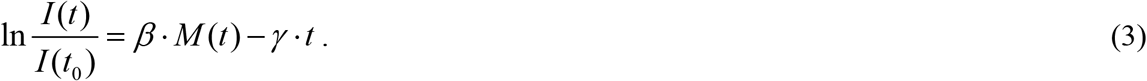

Or,

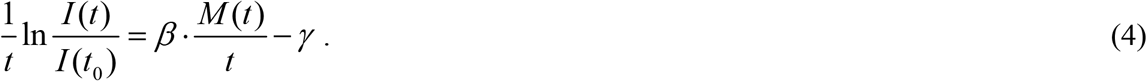

For a given community with a given reference time, the locus of 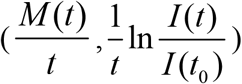 should be on a straight line, from which both *β* and *γ* can be determined.

## Results and Discussions

To critically examine the aforementioned mobility-informed disease-transmission model using Eq. (4), we chose twenty counties in the United States with the highest COVID-19 fatalities as of July 11, 2020, which is listed in Table S1 of the Supporting Information. We need both the time series of current infectious population, as well as the time series of mobility for each county. Regarding the infectious population, the number of confirmed cases cannot be used directly due to the fact that a significant fraction of COVID-19 carriers are asymptomatic and undetected. Moreover, the COVID-19 testing capacity varies over time and location. Here, assuming that the mortality rate of COVID-19 does not vary over time without a collapsing healthcare system nor new effective pharmaceutical treatment, we chose instead to use the daily fatality to infer the infectious population in the past with a time delay. Thus, the infection ratio 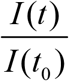 equals to 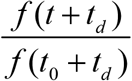 with *f(t)* being the daily fatality and *t*_*d*_ being the time delay. This conversion is valid as long as the mortality rate is constant over time for a given county even if it varies from county to county. The time delay *t*_*d*_ essentially is the average time from infection to death. For all counties, the time delay is 35 days with the only exception of New York City (New York, New York) with a time delay of 20 days. This time delay is reasonable considering it has been reported the time until hospitalization is five days and the mean hospitalization for critical care patient is 16 days according to Imperial College.^21^ New York City is unique due to its high population density with its hospitals nearing capacity at the height of the pandemic. The time-series of COVID-19 fatality used here was acquired on July 12, 2020 (data up to July 11, 2020), which is curated by Johns Hopkins University.^22^ To reduce fluctuation, the daily fatality is subjected to a running average over a span of five days.

Regarding the mobility, we resort to Google “COVID-19 Community Mobility Reports”^23^ which contain the changes in visits to places (i.e., activity locations), relative to a baseline of five-week average between January 3 to February 6, 2020. The places are divided into six different categories. For convenience, the “retail and recreation” mobility is used here to represent the overall mobility. As the Google mobility report is per day, the integral mobility *M(t)* here is essentially the cumulative mobility from a reference date. The Google mobility data was also acquired on July 12, which contains data up to July 7, 2020.

Figure 2 shows 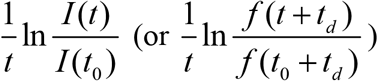 as a function of 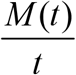 for twenty hardest-hit counties in the United States from a reference date (five-day running-average first reaching 1.5) to July 11, 2020 for the fatality data. The reference date for all twenty counties can be found in Table S1. The first twenty days after the reference date are not shown as data points are scattered at the onset of an outbreak. It is clear that all twenty counties show roughly a linear plot. The R^2^ value of the linear fitting is listed in Table S1. Hudson in New Jersey shows the lowest R^2^ value which is probably related to its uncommon trend of daily fatality. The model works particularly well for New York City, which shows almost a perfect linear line. It should be noted that the raw fatality and mobility data are used here, without removing outlier points (such as the fatality spikes in a number of New Jersey counties, as seen in Figure 3).

**Figure 2.**
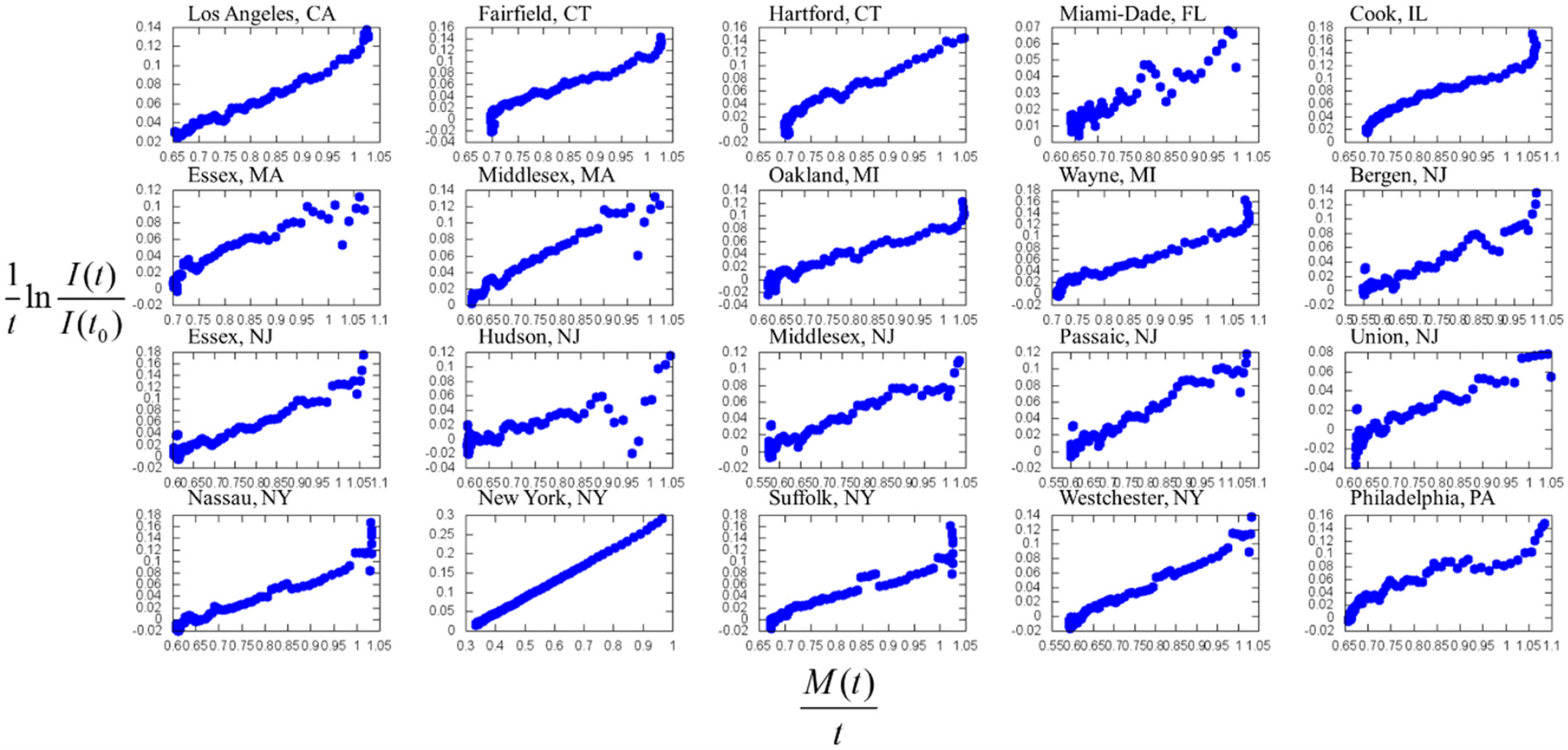
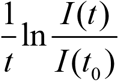 (time averaged natural logarithmic of relative infection) is plotted against 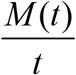 (average cumulative mobility) for twenty hardest-hit counties in the United States. Most counties show a linear relation, as expected from Eq. (4). We use Google mobility in the category of “retail and recreation”, relative to a baseline value. The infectious population ratio is inferred from the fatality ratio with a delay time of 35 days (except New York City, for which the delay time is 20 days). The reference date *t*_*0*_ is selected as the five-day running average fatality reaches 1.5. The first twenty days of data are not shown due to significant noise. The daily fatality is up to July 11, 2020.

**Figure 3.**
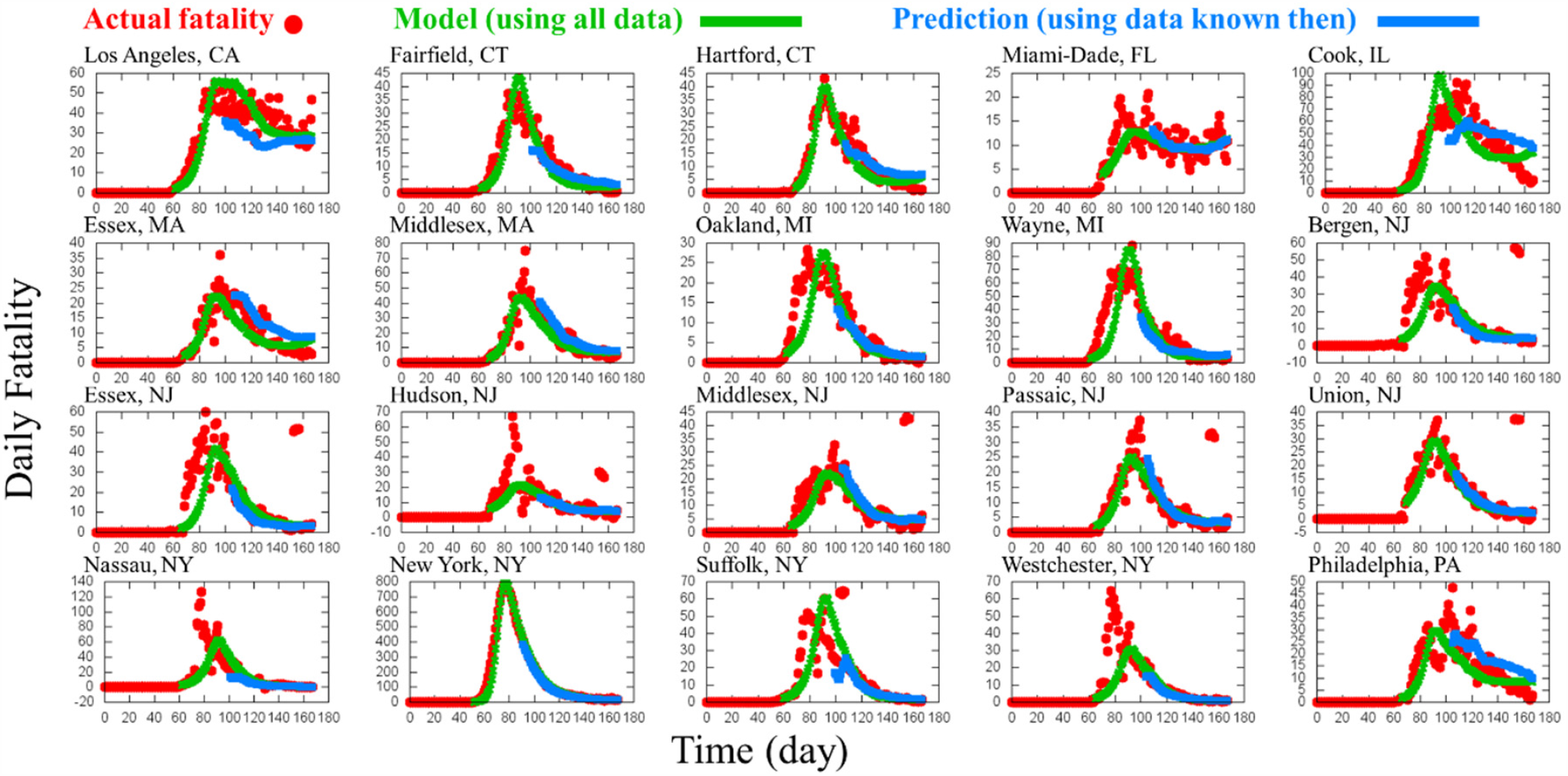
Daily fatality (red dots) for twenty hardest-hit counties in the United States. Time origin is January 22, 2020. The last day is July 11, 2020. The green lines are calculated according to Eq. (2) with the infection rate and recovery rate obtained from Figure 2, including all data available. The blue lines are also calculated according to Eq. (2), yet each fatality data point of a particular date is using slightly different parameters obtained from fitting data up to that date. Therefore, the blue line is the prediction using only information available then. The blue line prediction starts at 40 days after the reference date *t*_*0*_.

The infection rate *β* and recovery rate *γ* can be conveniently estimated from Figure 2, tabulated in Table S1. Based on the input data, both the infection rate and the recovery rate have a unit of *day*^-1^. The average infection rate over all twenty counties is around 0.3 *day*^-1^, which means: if people behave normally (same mobility as the baseline), without recovery, COVID-19 cases will increase 30% every day (i.e., double roughly every three days). The average recovery rate is around 0.16 *day*^-1^. Under normal mobility, the corresponding basic reproduction number is ∼1.9. From the recovery rate, one can infer an average recovery time of about 6 days. The recovery time is reasonable as it has been shown that infectious virus can be isolated from patient samples in the first eight days after illness but not afterwards.^24^ In addition, the mean serial interval between clinical onsets was estimated to be 5.8 days among 94 patients in China.^25^ With county-specific infection rate and recovery rate, one can use Eq. (2) to calculate the current infected (or daily fatality in the future) based on the cumulative mobility using infection rate and recovery rate in Table S1, as shown in Figure 3 (green lines). The model reproduces the daily fatality trend from the mobility data reasonably well. Figure 3 (blue lines) also shows the 35-day fatality prediction (20-day prediction for New York City) based on the information known then. In other words, the infection rate and recovery rate are different from Table S1 and are updating daily. The fatality prediction agrees reasonably well with the actual fatality.

It is also instrumental to visualize the daily fatality and cumulative mobility of all counties together to further understand the model. Recast Eq. (2) as,

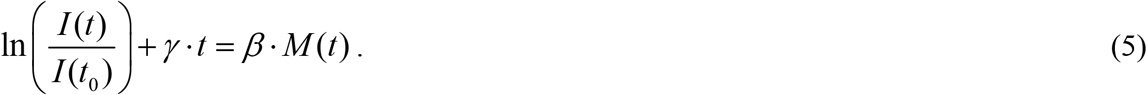

Now we can plot ln 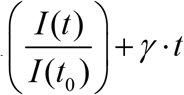 as a function of *β* · *M* (*t*) for all twenty counties, shown in Figure 4.

**Figure 4.**
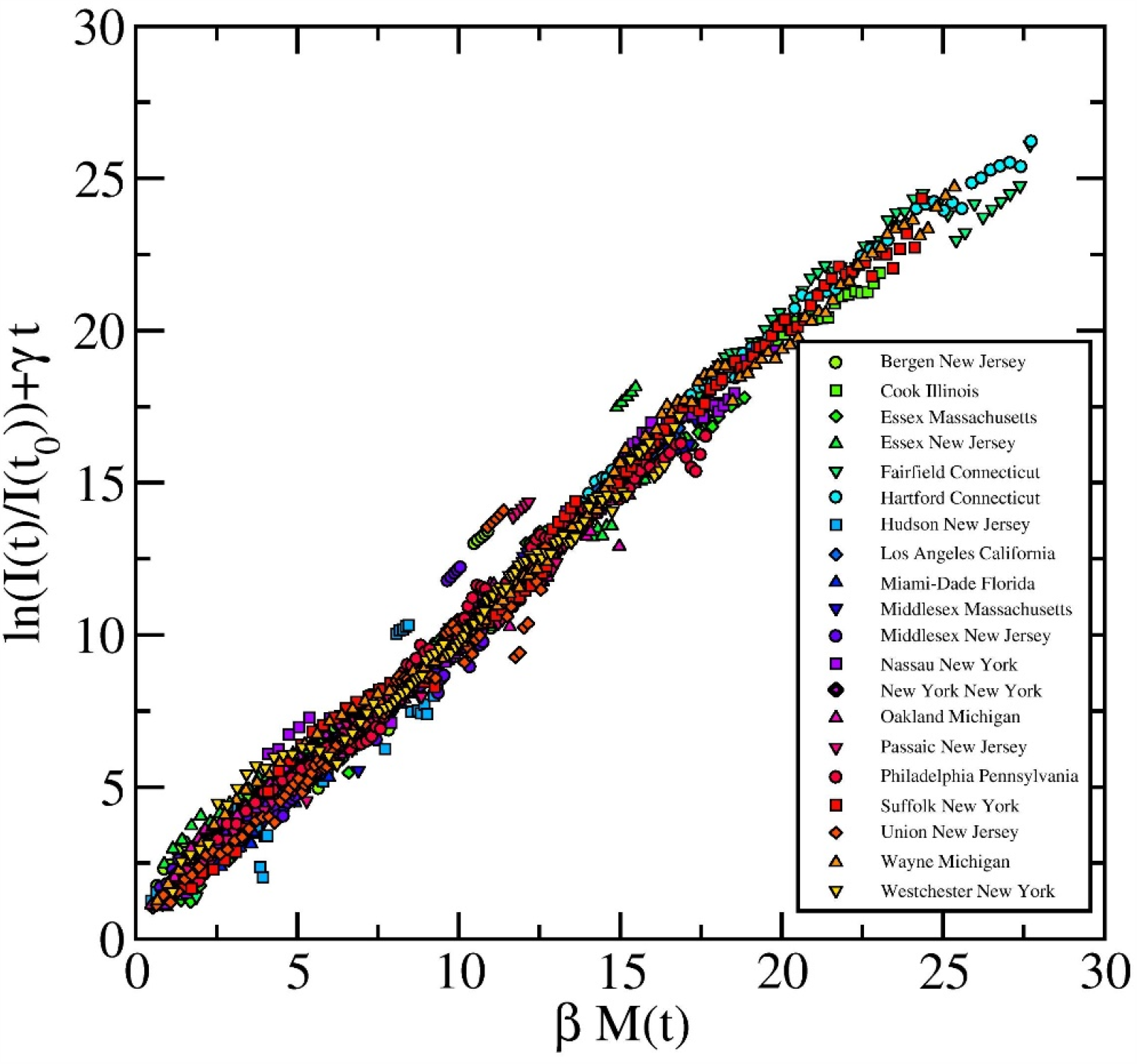
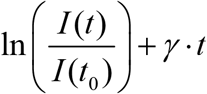 is plotted as a function of *β* · *M* (*t*) for all twenty hardest-hit counties in the United States. The infectious population data and mobility data are the same as in Figure 2. The infection and recovery rates for each county are obtained from linear fitting in Figure 2. Data points from all counties collapse into a linear line with a slope of 1 passing the origin, as expected from Eq. (5).

As expected, data points from different counties collapse into a linear line with a slope of 1 passing the origin. Figure 4 essentially shows how COVID-19 infection will grow without any recovery, as a function of cumulative mobility, which is indeed exponential. Importantly, the cumulative mobility plays the role of time in dictating the COVID-19 outbreak. Therefore, the obvious non-medical strategy of fighting COVID-19 pandemic is to control the cumulative mobility for every community.

The mobility-informed disease-transmission model offers a quantitative guideline in terms of mobility. Let us go back to Eq. (1), from which a critical mobility *m*_*c*_ can be defined,

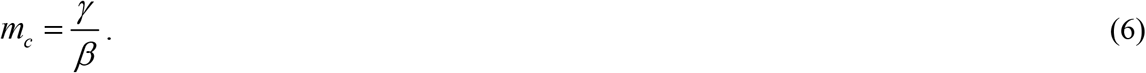

If the current mobility is higher than *m*_*c*_, new infection outpaces recovery, resulting in outbreak, and vice versa. It is interesting to note that the critical mobility is analogous to the reciprocal of the basic reproduction number *R*_*0*_, yet with very different model assumptions and physical meaning of the infection rate. More importantly, the basic reproduction number is obtained without mobility information which varies over time, while the critical mobility is obtained using mobility information and does not vary for a community (unless strict face-mask mandate is imposed). Figure 5 plots the critical mobility against the population density for twenty counties. It appears that the critical mobility centered around 60% for all counties, except the New York City (at ∼30%), which is not surprising due to its population density. The critical mobility seems to not vary much if a different delay time is chosen, as demonstrated for New York City and Los Angeles in Figure S4. Figure 5 also shows the month-average mobility from April to July 2020, which is consistent with the recent resurgence of COVID-19 cases especially in some of the southern states.

**Figure 5.**
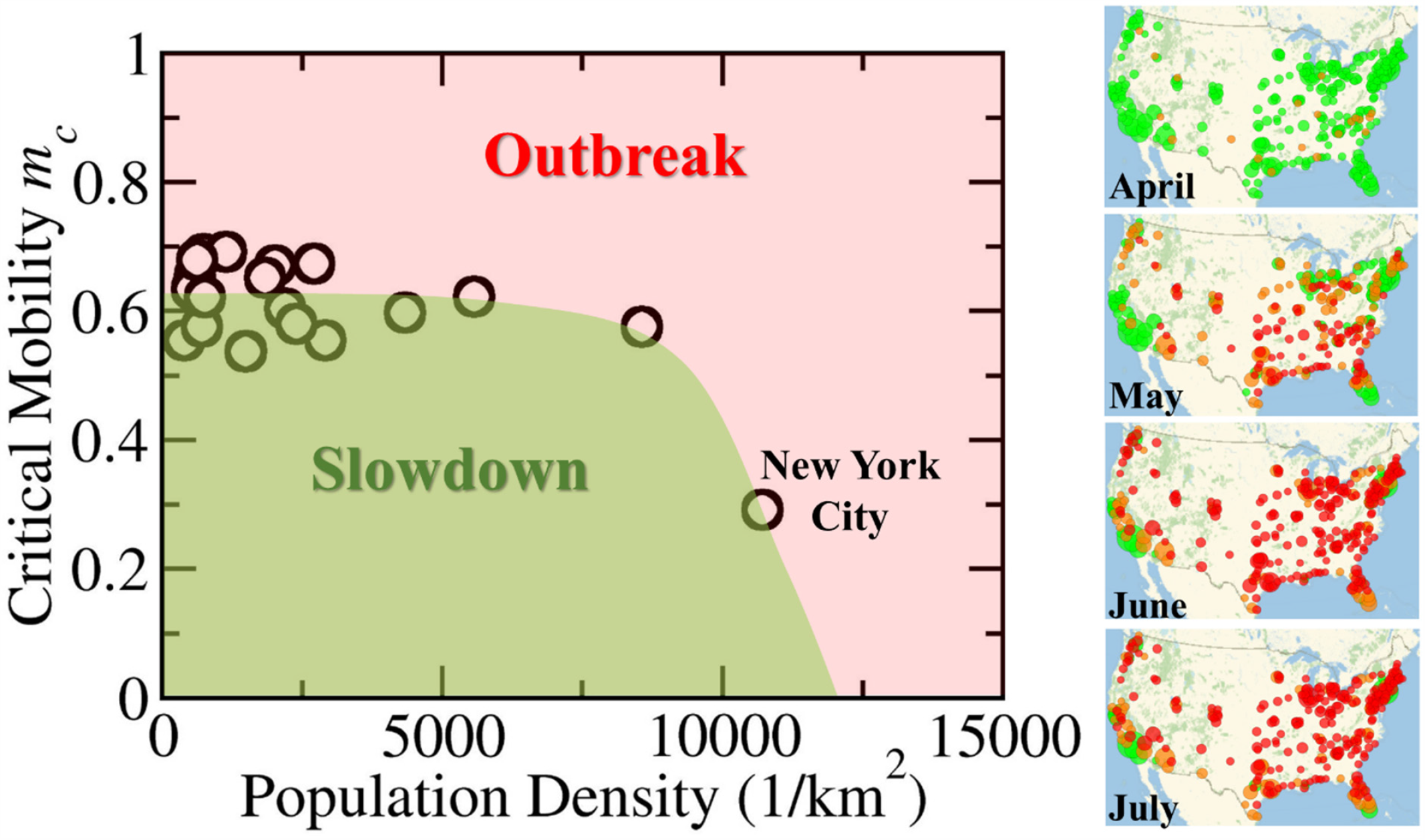
Critical daily mobility *m*_*c*_ is plotted as a function of population density for the twenty hardest-hit counties in the United States. It appears that most counties have a roughly constant *m*_*c*_ of around 60%, while *m*_*c*_ of New York City is around 30%. Depending on the daily mobility, each community faces either outbreak (daily mobility is higher than *m*_*c*_) or slowdown (daily mobility is lower than *m*_*c*_). We also plot the average mobility for about 325 counties with populations higher than 200,000 from April to July (only about one week of mobility is available for July), each colored green (lower than 70%), orange (70% to 80%), and red (over 80%), with the disk area proportional to the population. The increasing mobility in May to July shown in the figure is consistent with the drastically increasing number of cases (i.e., outbreaks) especially in some of the southern states.

The obvious message for policy makers is to utilize the critical mobility value to cap the daily mobility in monitoring reopening. As the results presented so far uses only the “retail and recreation” category of Google mobility, we also examined the “grocery and pharmacy”, “transit station” and “workplace” mobility. The other two categories are not included as the “residential” mobility varies generally in the opposite direction with the overall mobility, while the “parks” mobility has more complex behavior depending on the location. As shown in Figure S2 and S3, “transit station” and “workplace” work almost as well as “retail and recreation”, while “grocery and pharmacy” performs slightly worse. The critical mobility for different categories are given in Figure S3, such that the mobility of “transit station” should be capped around 50%, the mobility of “retail and recreation” and “workplace” should be capped around 60%. The critical mobility of “grocery and pharmacy” is around 90% which indicates it plays a lesser role in COVID-19 transmission. One could aggregate a composite weighted mobility from the above four types of data to better quantify mobility. We envision that the critical mobility using such a composite mobility will lie in-between the calculated critical mobility from individual types of mobility as shown in Figure S3.

In addition to monitor daily mobility in reference to the critical mobility, the second message for policy makers is to use the average mobility to help devise reopening plan or impose stay-at-home orders. The key insight is that the average mobility over a period of time must be lower than the critical mobility *m*_*c*_ to control COVID-19. For instance, if the mobility of a county in the past 30 days is 80% (assuming its *m*_*c*_ is 60%), this county may need to impose a stay-at-home order for the next 15 days (or 30 days) if the mobility can be reduced to 20% (or 40%) to average out the excessive mobility.

Without considering explicitly the effects of public health intervention measures such as face masking, or changing of human behaviors over time, it is surprising to see this model works well using solely the mobility data. This is probably due to the fact that all large gatherings (schools, Universities, sports, concerts and political rallies) were mostly closed for the duration of the data used in this model, which could have very different transmission characteristics due to high local population density. In addition, the late adoption of mask mandates, lack of effective enforcement, and general reluctance in following mandates by the citizens, all contribute to a roughly constant infection rate for each community. Due to the recent second wave of COVID-19, more states and local governments move to mask mandates, a time-dependent infection rate may be needed to apply the model to data of coming months. An expected consequence is that the critical daily mobility *m*_*c*_ will increase accordingly. For instance, let us assume that a face-mask mandate reduces 35% of the COVID-19 transmission compared to the pre-mandate period, the critical daily mobility will increase from a pre-mandate 60% to over 90%. Therefore, an effective face-mask mandate could raise the critical mobility back to normal level. For communities in which the mobility is difficult to regulate, face-mask mandate is necessary. On the other hand, for communities in which enforcing masks is not practical or popular, local government can choose to cap the daily mobility as a convenient tool to control COVID-19 spreading.

## Conclusions

In summary, we developed a mobility-informed infectious disease model, based on the collision theory used in understanding the kinetics of gas-phase chemical reactions. This simplified model yields an analytical solution to the infectious population, with the cumulative mobility dominating in the exponent thus controlling the disease transmission. Using fatality data from late January to early July to infer infectious population, as well as mobility data from mid-February to early July, we show the COVID-19 transmission in twenty hardest-hit counties of the United States agrees well with this model. From the fitting parameters obtained from all twenty counties, it appears that the critical daily relative mobility is around 60% (except the New York City, which is around 30%). This information can be utilized by different levels of governments to regulate reopening, particularly in the absence of mask mandates or enforcements.

## Data Availability

We use data publicly available from Google and JHU (Github).

## Acknowledgements

We thank invaluable discussions with Dr. Yanfang Su and Dr. Ka-Kit Tung from the University of Washington, and Dr. Liping Huang from Rensselaer Polytechnic Institute. We would also like to thank Dr. Qiran Xiao from Apple Inc. for insightful tips in data pre-processing.

## Supporting Information

**Table 1.** Important model information for the twenty hardest-hit counties in the United States in terms of the reference date (five-day running average daily fatality reaches 1.5), the infection rate *β*, recovery rate *γ* and critical daily mobility *m*_*c*_. The parameters are obtained using all data up to July 12, 2020.

| State | County | Population Density (1/km<sup>2</sup>) | Reference Date $t_0$ | $t_d$ (day) | $\beta$ (1/day) | $\gamma$ (1/day) | $R^2$ | $m_c$ |
| --- | --- | --- | --- | --- | --- | --- | --- | --- |
| California | Los Angeles | 2910 | 3/22/2020 | 35 | 0.26 | 0.14 | 0.98 | 55% |
| Connecticut | Fairfield | 744 | 3/23/2020 | 35 | 0.37 | 0.26 | 0.94 | 69% |
| Connecticut | Hartford | 2713 | 3/30/2020 | 35 | 0.40 | 0.27 | 0.95 | 67% |
| Florida | Miami-Dade | 400 | 3/31/2020 | 35 | 0.13 | 0.07 | 0.87 | 55% |
| Illinois | Cook | 2195 | 3/21/2020 | 35 | 0.31 | 0.18 | 0.96 | 60% |
| Massachusetts | Essex | 582.6 | 3/29/2020 | 35 | 0.27 | 0.17 | 0.90 | 65% |
| Massachusetts | Middlesex | 709.6 | 3/29/2020 | 35 | 0.29 | 0.17 | 0.94 | 58% |
| Michigan | Oakland | 535 | 3/23/2020 | 35 | 0.24 | 0.15 | 0.96 | 63% |
| Michigan | Wayne | 1148 | 3/22/2020 | 35 | 0.33 | 0.23 | 0.96 | 69% |
| New Jersey | Bergen | 1499.8 | 3/27/2020 | 35 | 0.21 | 0.12 | 0.90 | 54% |
| New Jersey | Essex | 2398 | 3/27/2020 | 35 | 0.28 | 0.16 | 0.93 | 58% |
| New Jersey | Hudson | 5566 | 3/30/2020 | 35 | 0.16 | 0.10 | 0.61 | 62% |
| New Jersey | Middlesex | 1496 | 3/27/2020 | 35 | 0.19 | 0.10 | 0.90 | 54% |
| New Jersey | Passaic | 8563.6 | 3/27/2020 | 35 | 0.22 | 0.13 | 0.93 | 58% |
| New Jersey | Union | 2014 | 3/30/2020 | 35 | 0.20 | 0.14 | 0.84 | 67% |
| New York | Nassau | 1840 | 3/23/2020 | 35 | 0.31 | 0.20 | 0.95 | 65% |
| New York | New York | 10715 | 3/13/2020 | 20 | 0.42 | 0.12 | 1.00 | 29% |
| New York | Suffolk | 632 | 3/21/2020 | 35 | 0.34 | 0.23 | 0.93 | 68% |
| New York | Westchester | 760 | 3/26/2020 | 35 | 0.28 | 0.17 | 0.98 | 62% |
| Pennsylvania | Philadelphia | 4337 | 3/27/2020 | 35 | 0.26 | 0.16 | 0.90 | 60% |

**Figure S1.**
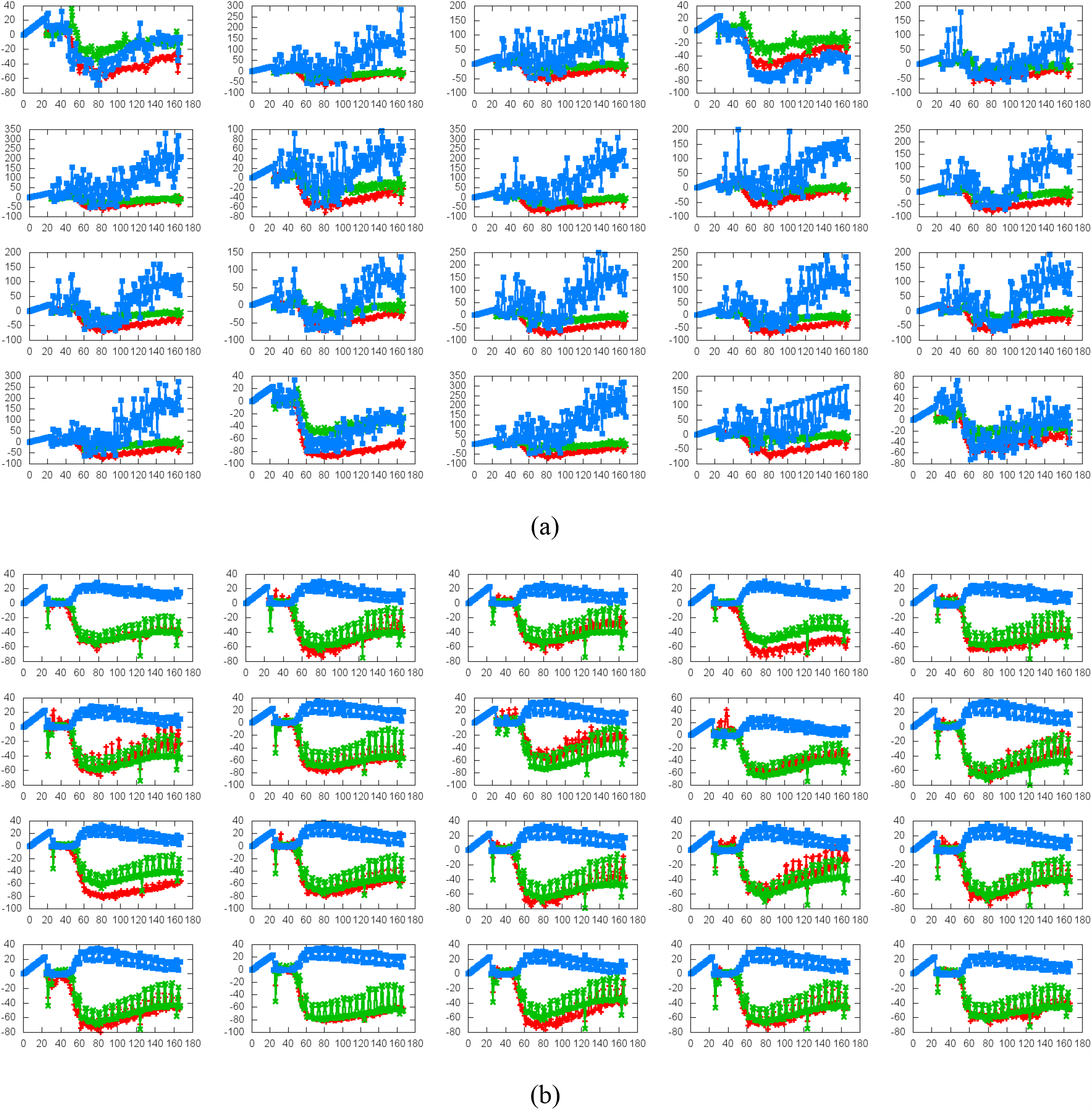
Google mobility as a function of time for the twenty hardest-hit counties in the United States. Time origin is January 22, 2020. (a) Retail and recreation (red), grocery and pharmacy (green) and parks (blue). (b) Transit station (red), workplace (green) and residential (blue). Parks and residential mobility behave very differently from the rest four categories of mobility. Note that the raw data from google mobility is plotted here. Relative mobility *m* used in our model is calculated from raw data *x* by *m = (100 + x)/100*.

**Figure S2.**
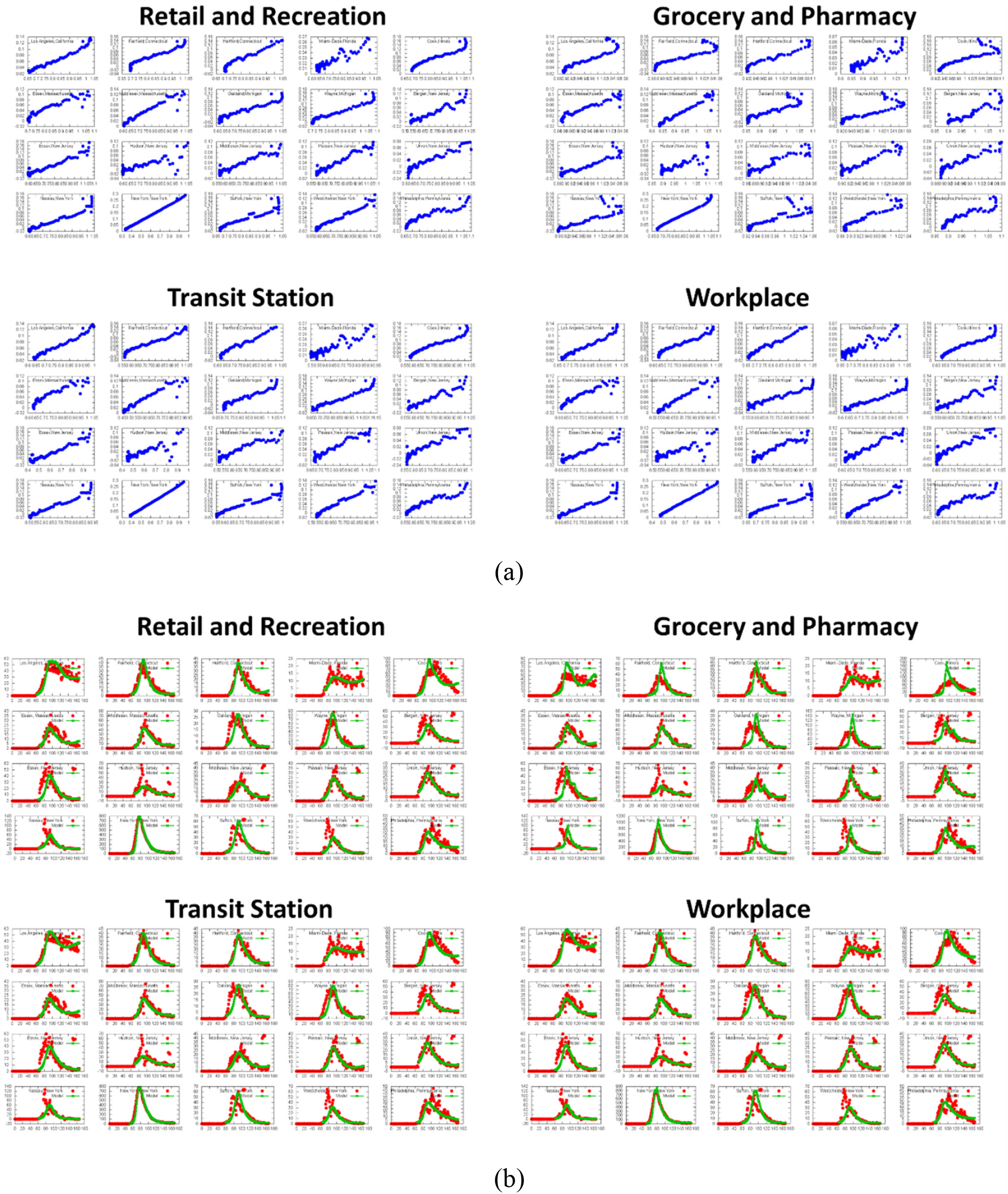
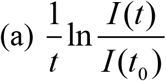 (time averaged natural logarithmic of relative infection) is plotted against 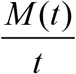 (average cumulative mobility) for the twenty hardest-hit counties in the United States using different categories of Google mobility as labeled. The one using retail and recreation mobility is the same plot as shown in Figure 2. (b) Daily fatality (red dots) and calculated fatality (green lines) using different categories of Google mobility. The one using retail and recreation is the same plot as shown in Figure 3 without the prediction.

**Figure S3.**
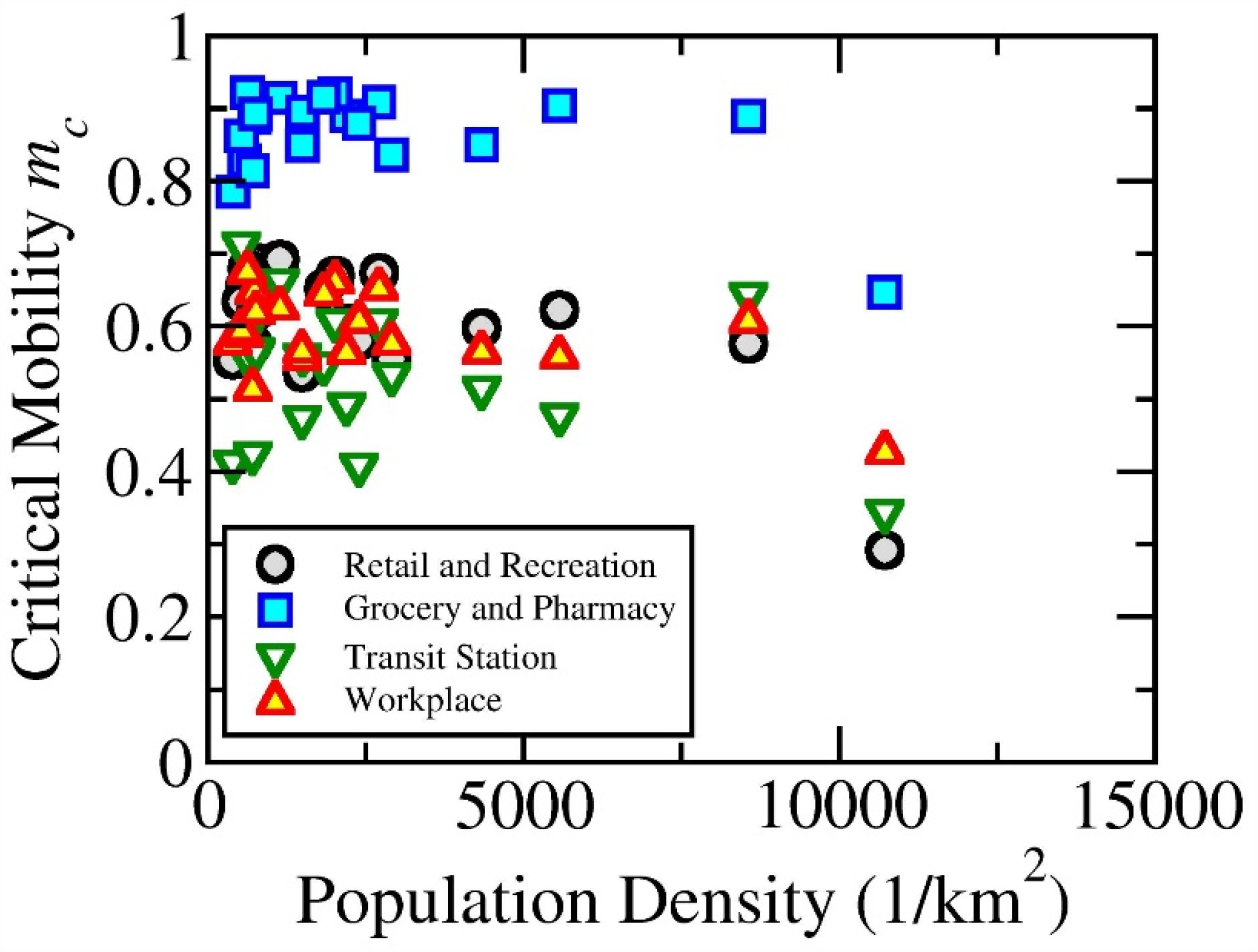
Critical daily mobility *m*_*c*_ is plotted as a function of population density for the twenty hardest-hit counties in the United States based on different categories of Google mobility. The critical daily mobility using the retail and creation mobility is the same as shown in Figure 5.

**Figure S4.**
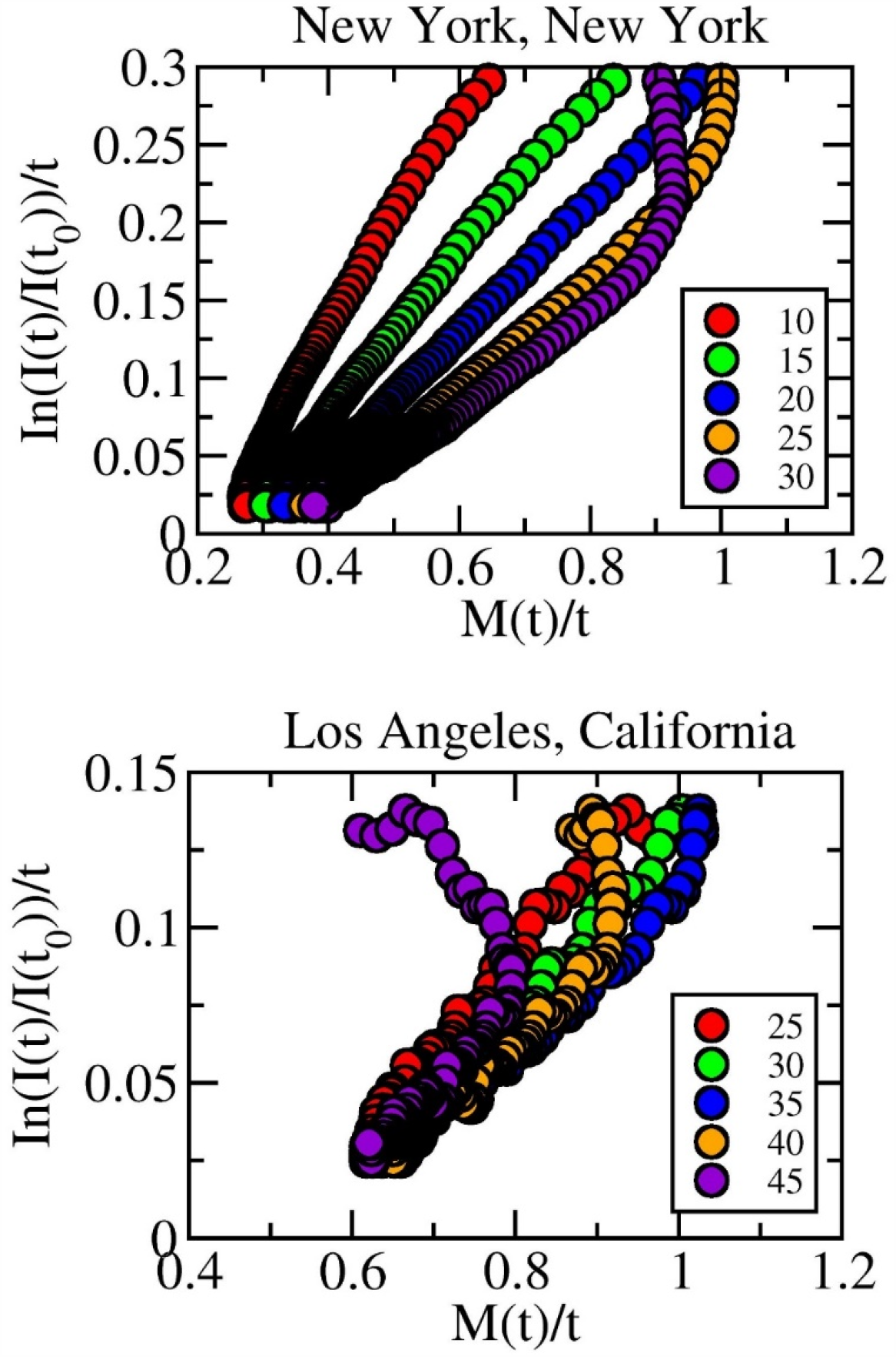
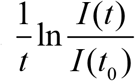 (time averaged natural logarithmic of relative infection) is plotted against 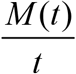 (average cumulative mobility) for New York city and Los Angeles, with different time delay *t*_*d*_ in days. For New York City, the linearity gets worsened either below or above 20 days. Similar observation can be made for Los Angeles, with 30 or 35 days both work well. It is also interesting to see that, the critical mobility (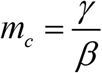, X-intercept) stays roughly the same for different values of time delay *t*_*d*_ for both New York City and Los Angeles, if fitting is applied to the linear portion.

## Notes

### Competing Interest Statement

The authors have declared no competing interest.

### Funding Statement

There is no funding support this work.

### Author Declarations

Not applicable to this work.

